# Trends in Fomepizole Use for Acetaminophen Poisoning in the United States; 2013-2024

**DOI:** 10.1101/2025.04.25.25326441

**Authors:** Mitchell D’Aloia, Dale Smith, Randy Boley, Emily Schamber, Dillon Thorpe, Trevonne M. Thompson, Neeraj Chhabra

## Abstract

**Background:** Fomepizole has been suggested as adjunctive therapy for severe acetaminophen poisoning though clinical efficacy is unknown. We sought to determine trends in the use of fomepizole for acetaminophen poisoning.

**Methods:** This is a cross-sectional analysis of hospitalized patients with acetaminophen poisoning from January 2013 through December 2024, using Epic Cosmos, a research database of 298 million patients nationally. We identified encounters involving acetaminophen poisoning by International Classification of Diseases, version 10 (ICD-10-CM) code. Data extracted included administration of N-acetylcysteine (NAC) and fomepizole, demographic data, and outcomes of death and liver transplantation. Data were analyzed using descriptive statistics to identify trends and multivariable logistic regression to determine associations with death or liver transplant.

**Results:** There were 114,111 hospital encounters involving acetaminophen poisoning with 64,957 (56.92%) receiving NAC, and 1,552 (1.36%) receiving fomepizole. In 2013, 0.44% of NAC-treated acetaminophen poisoning cases also received fomepizole. This rose to 6.27% in 2024. From 2013 to 2019, the proportion of NAC-treated acetaminophen cases receiving fomepizole was stable, but from 2019-2024, there was a 1029.64% increase in fomepizole use. Regression modeling indicated increased odds for death (OR=5.88, aOR=5.32 [95% CI: 4.52, 6.27]) and liver transplantation (OR=4.99, aOR=4.91 [95% CI: 2.10, 11.48]) among those who received fomepizole in addition to NAC, indicating increased fomepizole use in patients with severe toxicity.

**Conclusion:** Fomepizole use in acetaminophen poisoning has risen dramatically since 2019, particularly among patients at highest risk for death and liver transplantation. It is of critical importance to determine the efficacy of fomepizole for acetaminophen poisoning.

## Introduction

Acetaminophen is the most common cause of poisoning-related death reported to poison centers in the United States.^1^ Antidotal therapy for acetaminophen poisoning with N-acetylcysteine (NAC) is well established and effective. NAC treatment is nearly universally effective in preventing severe hepatotoxicity and death if initiated early and in sufficient quantity.^2,3^ Despite the presence of an effective focused intervention, morbidity and mortality from acetaminophen poisoning still occurs among patients seeking medical treatment, particularly among those that have already developed hepatotoxicity or those with large ingestions.^4^

Recently, fomepizole (4-methylpyrazole) has been suggested as adjunctive therapy for acetaminophen poisoning in cases with large ingestions or significant hepatotoxicity. This suggestion is largely based on in vitro and animal studies indicating inhibition of CYP2E1 metabolism of acetaminophen and c-Jun N-terminal kinase (JNK)-induced oxidant stress.^5,6^ Despite biological plausibility, clinical data on fomepizole use in acetaminophen poisoning is lacking. A recent consensus statement from a panel comprised of members of the leading clinical toxicology organizations in the United States and Canada concluded that the data available for fomepizole in acetaminophen poisoning “did not support a standard recommendation” on its use.^7^

Given the equipoise surrounding the use of fomepizole, we sought to determine national trends in fomepizole administration for acetaminophen poisoning. We hypothesized that fomepizole is being used with increasing frequency for acetaminophen poisoning and more commonly in cases of severe toxicity.

## Methods

This is a cross-sectional analysis of hospital admissions from January 1, 2013, to December 31, 2024, using the Epic Cosmos database. Epic Cosmos is a research system maintained by Epic Systems encompassing data from 1711 hospitals and 298 million patients and is broadly representative of demographics of the United States.^8^ We queried the Cosmos system for hospital admissions involving acetaminophen poisoning using a validated case definition from prior research based on International Classification of Diseases, Tenth revision, Clinical Modification (ICD-10-CM) code T39.1.^9^ Data extracted included administration of N-acetylcysteine (NAC), administration of fomepizole, demographic variables, and encounter outcomes including death and liver transplantation as indicated by ICD or Current Procedural Terminology (CPT) codes.

Data were analyzed using descriptive statistics to determine trends in fomepizole use by year. Not every case of suspected acetaminophen toxicity requires antidotal therapy. We therefore limited analyses of trends to patient encounters with significant acetaminophen poisoning as defined by hospital admission and receipt of NAC therapy. As the number of institutions contributing data to Cosmos varies by year, we report the use of fomepizole by year as a proportion of all encounters with significant acetaminophen poisoning, in addition to raw figures.

Each hospital admission with a corresponding ICD-10-CM code for acetaminophen poisoning was analyzed as an individual encounter, so an individual patient with multiple discrete hospital admission events for acetaminophen poisoning may have multiple encounters included in analysis. To determine associations between fomepizole use and death or liver transplantation, we performed multivariable logistic regression accounting for demographic variables including patient age, sex, race, ethnicity, and social vulnerability index (SVI).^10^ Laboratory results were not available within the Cosmos database at the time of analysis, so we were unable to control for degree of exposure or hepatotoxicity. Results are presented as adjusted odds ratios (aORs) with 95% confidence intervals. This study was deemed exempt as non-human subjects research by the primary institution. Analyses were performed in the Epic Cosmos system with visualizations in Python (v.3.11.7).

## Results

There were 114,111 encounters of acetaminophen poisoning among hospitalized patients identified from January 1, 2013, to December 31, 2024 (Table 1). Of these, 64,957 (56.92%) received NAC, and 1,552 (1.36%) received fomepizole. Among those meeting definition for significant acetaminophen poisoning in 2013, 0.44% (95% CI: 0.15, 0.72) received fomepizole. This rose to 6.27% (95% CI: 5.74, 6.80) in 2024. From 2013 to 2019, the proportion of significant acetaminophen poisoning cases receiving fomepizole increased only slightly (from 0.44% to 0.56%), but from 2019-2024, there was a 1019.64% increase in fomepizole use among patients with significant acetaminophen poisoning (Figure 1).

**Table 1.** Cohort Characteristics of Patients with Acetaminophen Poisoning.

| Variable | Hospitalized patients<br>n=114,111 | Cohort receiving NAC<br>n=64,957 | Cohort receiving NAC and fomepizole<br>n=1,501 |
| --- | --- | --- | --- |
| Age, median (IQR) | 41.45 (24 – 58) | 39.57 (23 – 54) | 44.35 (26 – 61) |
| Sex |  |  |  |
| Female | 79,415 | 45,489 | 1,031 |
| Male | 34,696 | 19,468 | 470 |
| Race |  |  |  |
| American Indian or Alaska Native | 2,564 | 1,550 | 31 |
| Asian | 3,452 | 2,148 | 50 |
| Black | 19,337 | 10,434 | 247 |
| Native Hawaiian or Other Pacific Islander | 457 | 290 | 6 |
| White | 81,252 | 46,320 | 1084 |
| Unknown/Other | 7,049 | 4,215 | 83 |
| Ethnicity |  |  |  |
| Hispanic | 12,874 | 7,529 | 165 |
| Non-Hispanic | 101,237 | 57,428 | 1,336 |
| Social Vulnerability Index |  |  |  |
| Lowest quartile | 28,759 | 16,469 | 359 |
| 2nd quartile | 26,490 | 15,171 | 354 |
| 3rd quartile | 27,450 | 15,559 | 360 |
| 4th quartile | 30,283 | 17,051 | 405 |
| Unknown | 1,129 | 707 | 13 |

**Figure 1.**
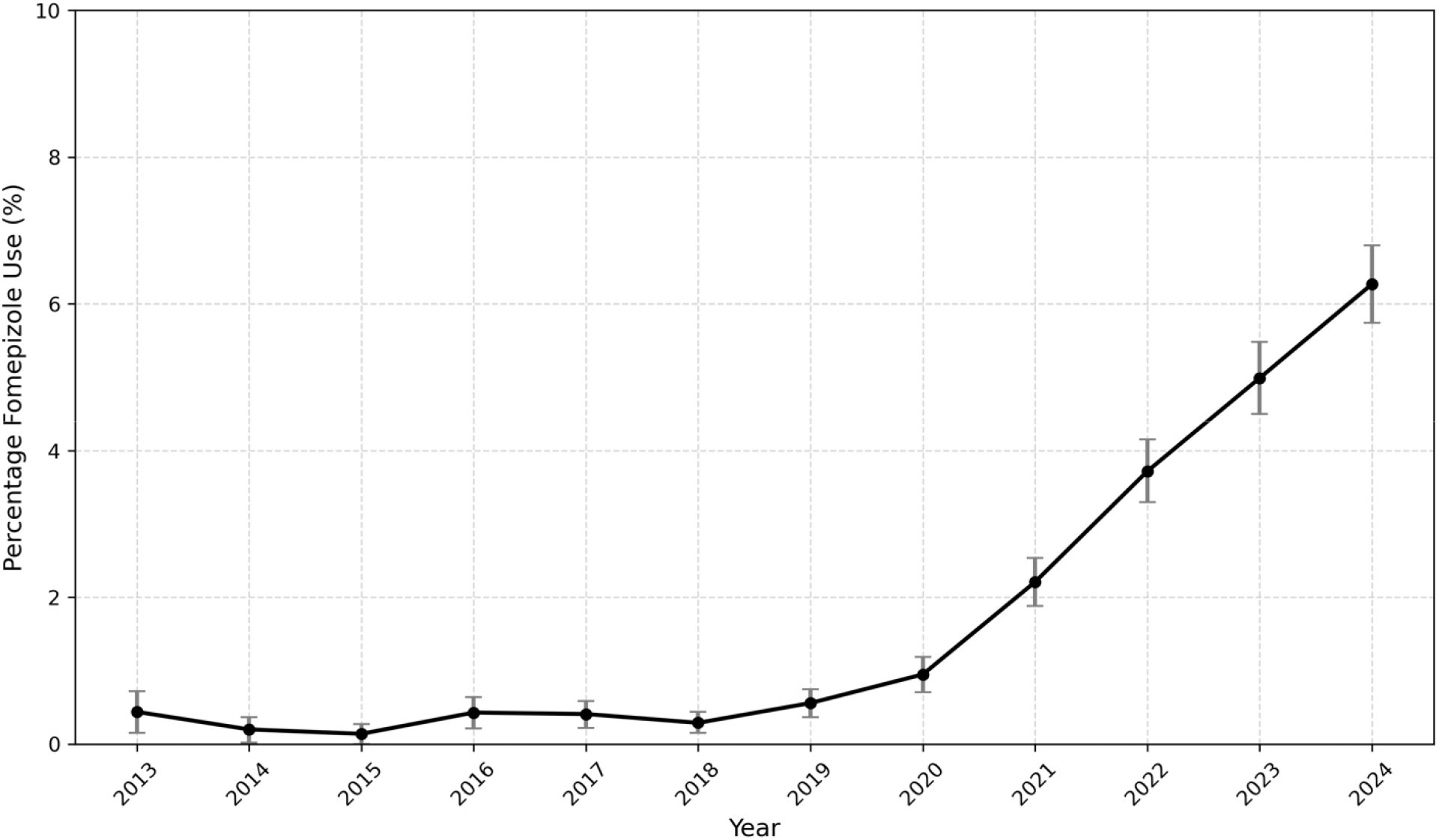
Trends in fomepizole use by year in patients with significant acetaminophen toxicity. *Significant acetaminophen toxicity defined as those requiring hospital admission and treatment with N-acetylcysteine

In regression modeling, the odds for death (OR=5.88, aOR=5.32 [95% CI: 4.52, 6.27], p<0.001) and liver transplantation (OR=4.99, aOR=4.91 [95% CI: 2.10, 11.48], p<0.001) were higher among those who received the combination of NAC and fomepizole when compared to those who received NAC alone. No demographics were significant predictors of transplant in adjustment models (all ps > .327), though small cell sizes and potentially low power should be considered in this result. Death was more common among older, male, and non-Hispanic patients, though the only race difference compared to White patients was a reduction in the odds of death among Asian patients (Table 2).

**Table 2.** Logistic regression results predicting death.

| Predictor | Death |  |
| --- | --- | --- |
|  | OR (95% CI) | P |
| Both fomepizole and NAC vs NAC alone | 5.32 (4.52, 6.27) | < .001 |
| Sex (ref: Female) | 1.14 (1.03, 1.26) | .010 |
| Ethnicity (ref: Hispanic) | 1.40 (1.13, 1.74) | .002 |
| Race (ref: White) |  |  |
| American Indian & Alaska Native | 0.91 (0.61, 1.34) | .624 |
| Asian | 0.69 (0.49, 0.99) | .045 |
| Black | 1.07 (0.93, 1.23) | .362 |
| Native Hawaiian or Pacific Islander | 0.70 (0.26, 1.91) | .492 |
| Other | 1.27 (0.94, 1.73) | .120 |
| Age | 1.04 (1.04, 1.04) | < .001 |
| SVI | 1.51 (1.28, 1.79) | < .001 |

SVI: Social Vulnerability Index

## Discussion

To our knowledge, this is the first study to examine trends in the use of fomepizole for acetaminophen poisoning using a large national database. Our results highlight a precipitous rise in the use of fomepizole for significant acetaminophen poisoning beginning in 2019. We also show that the use of fomepizole is highly associated with increased odds for both death and liver transplantation.

We urge caution in the interpretation of the association of fomepizole administration with negative clinical outcomes as causation. It is likely these associations represent increased fomepizole use in patients with more severe toxicity, rather than direct harm from the medication.

This interpretation is supported by surveys among medical toxicology subspecialists and a previous case series of 25 patients indicating higher likelihood to administer fomepizole in cases of severe poisoning.^11,12^ This suggests that fomepizole is being primarily employed in the sickest subset of patients.

The rise in fomepizole use for acetaminophen poisoning is despite the lack of human trials indicating a direct clinical benefit of the intervention.^6,7^ Both in vitro and animal studies have suggested a potential therapeutic benefit for fomepizole in humans with acetaminophen toxicity primarily through two mechanisms: inhibition of CYP2E1 which metabolizes acetaminophen to toxic metabolite N-acetyl-p-benzoquinone imine, and inhibition of JNK which prevents oxidative stress and apoptosis in affected organs. Given the rising use of fomepizole, particularly among the sickest patients, it is critical that high-quality clinical and translational research identify what, if any, benefit there is to the medication and, if benefit exits, which patients are most likely to benefit.

These results should be interpreted within the context of the study’s limitations. As a retrospective study using data collected for non-research purposes, some data may be missing or inaccurate, such as those used to determine the type of exposure, degree of exposure, and whether consultation with a poison center or medical toxicology subspecialist was sought. Similarly, the lack of laboratory data available at the time of analysis in the Epic Cosmos system limits evaluation of degree of hepatotoxicity for selected cases. While the database is broadly representative of the US population, it may not perfectly represent all hospitalized patient encounters. Our evaluation relies upon ICD-10-CM codes for the identification of cases of acetaminophen poisoning. While a validated approach with 90% sensitivity, some cases of acetaminophen poisoning were likely excluded due to under coding.^9^

## Conclusion

Fomepizole use is increasing in frequency and being used to treat the most severe cases of acetaminophen poisoning despite a lack of clinical evidence in humans supporting its use. Further studies are needed to characterize its clinical benefit and determine its utility in the treatment of acetaminophen poisoning.

## Data Availability

The data that support the findings of this study are available from Epic Cosmos. Restrictions apply to the availability of these data, which were used under license for this study. Data are available from https://cosmos.epic.com with the permission of Epic Cosmos.

## Acknowledgment

Data used in this study came from Epic Cosmos, a dataset created in collaboration with a community of Epic health systems representing more than 298 million patient records from over 1711 hospitals and 39.9 thousand clinics from all 50 states, D.C., Lebanon, and Saudi Arabia.

This research was presented as an abstract at the American College of Medical Toxicology (ACMT) Annual Scientific Meeting in April 2025 in Vancouver, BC, Canada.

